# Evaluation of the risk relationship between average alcohol volume consumed and suicide: An analysis of mortality linked cohort data

**DOI:** 10.1101/2023.11.22.23298895

**Authors:** Shannon Lange, Yachen Zhu, Charlotte Probst

## Abstract

**Objective:** To evaluate the relationship between average alcohol volume consumed per day and suicide.

**Methods:** Data from the annual, cross-sectional US National Health Interview Survey, 1997-2018, was obtained, and linked to the 2019 National Death Index. The association between average alcohol volume consumed in grams per day (g/day) and suicide was quantified using Cox proportional hazards model (multiplicative) and Aalen’s additive hazard model. All analyses were stratified by sex, and adjusted for education, marital status, race/ethnicity, and survey year.

**Results:** On the multiplicative scale, for males, former drinkers and those who consumed on average (40, 60] g/day had about 53% (HR=1.53, 95% CI: 1.10, 2.13) and 77% (HR=1.77, 95% CI: 1.17, 2.66) greater risk of dying by suicide, compared to lifetime abstainers, respectively. There was no significant association found for former or current drinkers among females, on the multiplicative scale. On the additive scale, for males and females, being a former drinker was associated with 11.4 (95% CI: 2.3, 20.4) and 5.6 (95% CI: 0.8, 10.4) additional deaths per 100,000 person years, compared to lifetime abstainers. For males only, drinking (40, 60] g/day on average was associated with 23.2 (95% CI: 6.7, 39.7) additional deaths per 100,000 person years. Level of education was not found to modify the focal relationship for males or females.

**Conclusions:** The findings suggest that the relationship between average alcohol volume consumed per day and suicide is nuanced. Additional research on the respective relationship is needed, including repeated measures of average alcohol consumption over time.

**What is already known on this topic:** There is a dearth of studies on the sex-specific relationship between average alcohol volume consumed per day and suicide. The one existing study, from South Korea, found that for males as average alcohol volume consumed increased, the likelihood of death by suicide also increased. For females it was not possible to estimate the risk associated with the upper level of consumption due to a zero-cell count.

**What this study adds:** This is the first systematic investigation of the sex-specific relationship between average alcohol volume consumed per day and death by suicide using a large linked dataset from the United States. It is also the first to evaluate the modifying effect of education, an important indicator of socioeconomic status, on the respective relationship.

**How this study might affect research, practice or policy:** The findings were not in line with the sparse existing literature, indicating that this line of research is not yet resolved and more research is needed.

## INTRODUCTION

Following nearly 20-years of steady increase (1), the suicide mortality rate in the United States (US) had decreased in 2019 and 2020 (2, 3). However, it appears that the suicide mortality rate in the US is once again on the rise (4, 5). Thus, innovative, evidence-based suicide prevention interventions are urgently needed to circumvent this trend. It is well-known that alcohol use is a risk factor for death by suicide (6), having both precipitating and predisposing effects (7, 8). However, much of the existing literature on the predisposing effects of alcohol focuses on alcohol use disorders (AUD) (9). Although such knowledge has suicide prevention implications, in that it is possible that AUD treatment is one prevention mechanism, the scope is limited. Estimating the relationship between average alcohol volume consumed per day and death by suicide would allow us to determine whether sub-clinical levels of drinking may also contribute to the risk of suicide, which may present another promising point of preventive intervention on a larger scale (e.g., via alcohol control policy).

Meaningful differences in the relationship between alcohol use and suicide across the sexes have been observed (10, 11), but again, this observation is largely limited to certain dimensions of alcohol use (i.e., acute alcohol use, and AUD). Based on a recent systematic review conducted by Lange and colleagues (12), only one study from South Korea (13) has provided sex-specific estimates of the relationship between average alcohol volume consumed and death by suicide to date. Thus, the vast majority of existing studies have either provided unstratified estimates, or have restricted their sample to males only. The study from South Korea by Jee and colleagues (13) indicates that as average alcohol volume consumed increases, the likelihood of death by suicide also increases. However, this study was limited to only two categories of average alcohol volume consumed and a risk estimate for females was not possible for the upper level, due to a zero-cell count.

In addition to being an important risk factor for death by suicide, alcohol has also been shown to contribute to socioeconomic inequalities in mortality (14). In fact, it has been shown that socioeconomic status (SES) acts as an effect modifier of the relationship between alcohol use and mortality, with increased risks of mortality for alcohol users with a low SES (15-17). However, there is a complete absence of studies investigating the effect modification between SES and alcohol use on the risk of death by suicide, representing a significant gap in knowledge. This gap in knowledge is particularly noteworthy for the US, as it is apparent that the suicide morality rate varies by level of education (18); an important indicator of SES.

Accordingly, the aim of the current study was to evaluate the sex-specific relationship between average alcohol volume consumed per day and death by suicide, and to explore whether level of education acts as an effect modifier of this relationship.

## METHODS

### Data

We obtained data from the annual, cross-sectional US National Health Interview Survey (NHIS), 1997-2018. NHIS collects health information from the civilian non-institutionalized US population residing in the 50 states and the District of Columbia. The average response rate for the NHIS 1997-2018 is 70%. The outcome was time to death by suicide or last presumed alive, which was based on linked data (i.e., NHIS data linked to the 2019 National Death Index (NDI)).

### Operationalizations

Death by suicide was operationalized using the International Classification of Diseases, 10^th^ revision (ICD-10) codes (X60-X84, Y87.0) and 9^th^ revision (ICD-9) codes (E950-E959). For each participant, the baseline age was calculated as the difference between the interview date and date of birth. The end age was calculated as the difference between date of death and date of birth if the participant was deceased by 12/31/2019, and as the difference between 12/31/2019 and date of birth if the participant was alive on 12/31/2019.

Average alcohol volume consumed in grams per day (g/day) was calculated by multiplying (the self-reported number of days participants drank alcohol in the past year divided by 365) and (the average number of drinks on days participants drank multiplied by 14 grams per drink), assuming 14 grams of pure alcohol per standard drink. We categorized alcohol intake in the past 12 months as: 1) lifetime abstainer (never drank alcohol in the past 12 months and never had 12+ drinks in any one year; reference group), 2) former drinker (never drank alcohol in the past 12 months but had 12+ drinks in any one year), 3) past year daily average of (0, 20] g for both males and females, 4) past year daily average of (20, 40] g for males and >20 g for females, 5) past year daily average of (40, 60] g for males only, and 6) past year daily average of >60 g for males only. The latter two categories were for males only, as there were relatively few females who consumed >40 g/day (those that had were combined with the former group, denoted as >20 g/day for females). Lifetime abstainers (rather than current abstainers) were used as the reference group to reduce the “sick-quitter” bias (i.e., the bias of categorizing individuals who have stopped drinking alcohol due to health concerns as the reference group) (19).

Covariates included education (high school diploma or less, some college, bachelor’s degree or more), marital status (married or living with partner vs never married, widowed, divorced, or separated), race and ethnicity (non-Hispanic Black, Hispanic, Others vs non-Hispanic White). The analyses were restricted to those 25 years of age and older at baseline, assuming that most of the participants had completed their highest level of education by then.

### Statistical analyses

We quantified the association between average alcohol volume consumed and death by suicide using both multiplicative and additive hazards regression models (20). We used age rather than follow-up years as the time scale to allow for a complete non-parametric age effect (21). All analyses were stratified by sex, and adjusted for education, marital status, race and ethnicity, and survey year. On the multiplicative scale, we applied Cox proportional hazards model to evaluate the association between alcohol use and time to death by suicide. We did not find any evidence for a violation of the proportional hazards assumption after evaluating the independence between Schoenfeld residuals and time for all covariates through statistical tests and graphical diagnostics (22). The complex survey design of the NHIS was accounted for in the Cox models using survey weights, strata, and primary sampling units. The Cox proportional hazards regression coefficients were exponentiated to obtain hazard ratios (HRs). On the additive scale, we used Aalen’s additive hazard model (23-26), a flexible semi-parametric model for survival analyses, to model the risk of death by suicide as a function of the explanatory variables plus an unspecified baseline risk. The effect estimates multiplied by 100,000 can be directly interpreted as the number of additional deaths by suicide associated with each risk factor per 100,000 person years (py). In contrast to the Cox proportional hazards model, the complex survey design of the NHIS cannot be accounted for in the Aalen’s models. An interaction effect of level of education and average alcohol volume consumed on death by suicide was tested in all models.

All statistical analyses were conducted in R, version 4.2.3. A two-sided p-value <0.05 was considered statistically significant.

## RESULTS

The overall sample was 562,632 (246,302 (47.7%, weighted) males and 316,330 (52.3%, weighted) females), with a mean age at baseline of 51.6 years (SD=16.9). Participants were followed an average of 10.7 years (SD=6.4). Overall, compared to females, males consumed more on average, regardless of education level. See Table 1 for participant characteristics at baseline, by sex and level of education.

**Table 1.** Participant characteristics at baseline, by sex and level of education.

|  | Overall | Males |  |  | Females |  |  |
| --- | --- | --- | --- | --- | --- | --- | --- |
|  |  | High school diploma or less | Some college | Bachelor's degree or more | High school diploma or less | Some college | Bachelor's degree or more |
| Sample size, n | 562,632 | 108,578 | 65,625 | 72,099 | 142,505 | 91,428 | 82,397 |
| Age at baseline, mean years (SD) | 51.57 (16.91) | 52.43 (16.86) | 49.21 (15.52) | 49.63 (15.82) | 55.79 (18.15) | 50.23 (16.51) | 48.18 (15.57) |
| Follow-up length, mean years (SD) | 10.67 (6.36) | 10.56 (6.34) | 10.54 (6.38) | 10.43 (6.37) | 10.97 (6.35) | 10.81 (6.35) | 10.47 (6.36) |
| Deaths by suicide, n (rate per 100,000 population [95% CI]) | 854 (13.7 [12.6, 14.9]) | 308 (25.1 [21.8, 28.4]) | 193 (23.9 [19.9, 27.9]) | 145 (17.6 [14.3, 21.0]) | 70 (4.2 [3.1, 5.2]) | 83 (7.6 [5.9, 9.3]) | 55 (6.8 [3.7, 9.9]) |
| Race/Ethnicity, n (%) |  |  |  |  |  |  |  |
| Non-Hispanic white | 368,305 (70.9) | 63,570 (64.2) | 46,486 (73.9) | 54,915 (78.5) | 80,842 (64.7) | 61,928 (73.3) | 60,564 (76.6) |
| Non-Hispanic black | 77,752 (11.2) | 15,937 (12.4) | 8,625 (11.5) | 5,489 (6.7) | 23,857 (13.3) | 15,156 (13.4) | 8,688 (8.5) |
| Hispanic | 87,242 (12.5) | 25,310 (20.0) | 7,655 (10.4) | 4,880 (5.9) | 32,375 (17.8) | 10,870 (9.4) | 6,152 (6.1) |
| Other | 29,333 (5.4) | 3,761 (3.5) | 2,859 (4.2) | 6,815 (8.9) | 5,431 (4.2) | 3,474 (3.9) | 6,993 (8.8) |
| Married/cohabitating, n (%) | 307,126 (67.9) | 64,195 (70.1) | 38,437 (70.8) | 46,240 (76.4) | 65,799 (59.6) | 45,363 (63.5) | 47,092 (70.0) |
| Alcohol use, mean grams per day (SD) | 5.49 (18.43) | 9.09 (29.32) | 8.92 (22.08) | 7.94 (18.00) | 2.24 (12.20) | 3.19 (9.82) | 4.02 (9.66) |
| Alcohol use, n (%) |  |  |  |  |  |  |  |
| Lifetime abstainer | 174,936 (29.0) | 28,869 (26.2) | 12,156 (18.4) | 12,081 (16.3) | 73,157 (49.0) | 29,036 (30.5) | 19,637 (23.1) |
| Former drinker | 40,482 (6.7) | 13,091 (11.1) | 5,688 (8.0) | 3,724 (4.8) | 9,425 (6.3) | 5,208 (5.4) | 3,346 (3.8) |
| (0, 20] g/day | 307,314 (57.0) | 53,084 (50.4) | 39,450 (61.5) | 48,263 (68.2) | 56,353 (42.1) | 54,009 (60.6) | 56,155 (69.2) |
| (20, 40] g/day (>20 g/day for females) | 28,292 (5.2) | 7,296 (6.7) | 5,096 (7.5) | 5,896 (8.0) | 3,570 (2.6) | 3,175 (3.5) | 3,259 (4.0) |
| (40, 60] g/day | 6,031 (1.1) | 2,906 (2.6) | 1,709 (2.5) | 1,416 (1.9) | n/a | n/a | n/a |
| >60 g/day | 5,577 (1.0) | 3,332 (3.0) | 1,526 (2.1) | 719 (0.9) | n/a | n/a | n/a |

n/a: not applicable
*Note.* All n's, means, and standard deviations are unweighted, rates per 100,000 population and proportions are weighted.

Overall, 854 individuals (646 males and 208 females) died by suicide during the follow-up period, for a suicide mortality rate of 13.7 (95% CI: 12.6, 14.9) per 100,000 py (males: 22.5 [95% CI 20.4, 24.5] per 100,000 py; females: 5.9 [95% CI 4.8, 7.0] per 100,000 py). Among males, the highest suicide mortality rate was among those with a high school diploma or less (25.1 [95% CI: 21.8, 28.4] per 100,000 py), followed by those with some college (23.9 [95% CI: 19.9, 27.9] per 100,000 py), and lastly those with a bachelor’s degree or higher (17.6 [95% CI: 14.3, 21.0] per 100,000 py). Among females, the highest suicide mortality rate was among those with some college (7.6 [95% CI: 5.9, 9.3] per 100,000 py), followed by those with a bachelor’s degree or higher (6.8 [95% CI: 3.7, 9.9] per 100,000 py), and lastly those with a high school diploma or less (4.2 [95% CI: 3.1, 5.2] per 100,000 py).

On the multiplicative scale, for males, former drinkers and those who consumed on average (40, 60] g/day had about 53% (HR = 1.53, 95% CI: 1.10, 2.13) and 77% (HR = 1.77, 95% CI: 1.17, 2.66) greater risk of dying by suicide, compared to lifetime abstainers, respectively (Table 2). On the additive scale, for males, being a former drinker or drinking (40, 60] g/day on average was associated with 11.4 (95% CI: 2.3, 20.4) and 23.2 (95% CI: 6.7, 39.7) additional deaths per 100,000 py, respectively. On a multiplicative scale, for females, compared to lifetime abstainers, former drinkers and current drinkers (regardless of amount consumed on average) did not have a statistically significantly greater risk of dying by suicide. However, for females, on the additive scale, being a former drinker was associated with 5.6 (95% CI: 0.8, 10.4) additional deaths per 100,000 py, compared to lifetime abstainers.

**Table 2.** Results of the hazards model^a^ evaluating the sex-specific relationship between average alcohol volume consumed and death by suicide.

|  | n | N | HR | 95% CI | p-value | Additional deaths<br>per 100,000<br>person years | 95% CI | p-value |
| --- | --- | --- | --- | --- | --- | --- | --- | --- |
| **Males** |  |  |  |  |  |  |  |  |
| Level of education |  |  |  |  |  |  |  |  |
| High school diploma or less | 308 | 108,578 | **1.39** | **1.11, 1.74** | **0.004** | **8.2** | **3.7, 12.6** | **<0.001** |
| Some college | 193 | 65,625 | **1.33** | **1.03, 1.71** | **0.027** | **8.4** | **3.5, 13.3** | **0.001** |
| Bachelor's degree or higher | 145 | 72,099 | Ref | - | - | Ref | - | - |
| Alcohol use |  |  |  |  |  |  |  |  |
| Lifetime abstainer | 125 | 53,106 | Ref | - | - | Ref | - | - |
| Former drinker | 82 | 22,503 | **1.53** | **1.10, 2.13** | **0.011** | **11.4** | **2.3, 20.4** | **0.014** |
| (0, 20] g/day | 334 | 140,797 | 0.91 | 0.71, 1.16 | 0.436 | -0.7 | -5.1, 3.8 | 0.769 |
| (20, 40] g/day | 51 | 18,288 | 1.07 | 0.75, 1.53 | 0.714 | 0.8 | -6.8, 8.5 | 0.836 |
| (40, 60] g/day | 32 | 6,031 | **1.77** | **1.17, 2.66** | **0.006** | **23.2** | **6.7, 39.7** | **0.006** |
| >60 g/day | 22 | 5,577 | 1.25 | 0.72, 2.16 | 0.429 | 10.3 | -4.8, 25.5 | 0.180 |
| **Women** |  |  |  |  |  |  |  |  |
| Level of education |  |  |  |  |  |  |  |  |
| High school diploma or less | 70 | 142,505 | 0.70 | 0.36, 1.35 | 0.286 | 0 | -2.2, 2.2 | 0.979 |
| Some college | 83 | 91,428 | 1.11 | 0.62, 1.99 | 0.732 | **2.6** | **0.1, 5.0** | **0.045** |
| Bachelor's degree or higher | 55 | 82,397 | Ref | - | - | Ref | - | - |
| Alcohol use |  |  |  |  |  |  |  |  |
| Lifetime abstainer | 48 | 121,830 | Ref | - | - | Ref | - | - |
| Former drinker | 19 | 17,979 | 1.66 | 0.80, 3.44 | 0.171 | **5.6** | **0.8, 10.4** | **0.022** |
| (0, 20] g/day | 129 | 166,517 | 1.13 | 0.59, 2.16 | 0.707 | 1.7 | 0.0, 3.4 | 0.052 |
| >20 g/day | 12 | 10,004 | 1.35 | 0.58, 3.16 | 0.489 | 6.0 | -0.6, 12.7 | 0.076 |

g: grams; HR: hazard ratio; Ref: reference category
<sup>a</sup>Adjusted for race/ethnicity, marital status, and survey year
Note. Bolded text signifies statistical significance at $\alpha=0.05$ .

Level of education was not found to modify the focal relationship for males or females in either the Cox proportional hazards model or Aalen’s additive hazard model (see Supplemental Material, Table S1).

## DISCUSSION

The current study is the first to generate detailed, high-quality evidence on the sex-specific risk of average alcohol volume consumed per day on death by suicide in the US. It was found that average alcohol volume consumed was not significantly associated with an increased risk of death by suicide among females; however, being a former drinker was associated with more than five additional deaths per 100,000 py. Among males, former drinkers and those who consumed an average of (40, 60] g/day had about 53% and 77% greater risk of dying by suicide and were associated with 11 and 23 additional deaths per 100,000 py, compared to lifetime abstainers, respectively. The finding that former drinkers had an increased risk of dying by suicide, compared to lifetime abstainers is consistent with the study among males in Japan by Akechi and colleagues (27), who found that the risk of suicide was 6.7-fold among ex-drinkers, compared with the risk in occasional drinkers (i.e., those who consumed alcohol 1-3 days per month). This elevated risk of death by suicide among former drinkers could reflect the health, economic, social and legal consequences that heavy alcohol use can have (28), which may have led to a desire to abstain, but that are also associated with an increased risk of suicide (e.g., see (29, 30)).

Further, although education level was not found to modify the relationship between average amount of alcohol consumed and death by suicide, lower levels of education (high school diploma or less and some college) were associated with an increased risk of suicide as well as additional deaths by suicide among males. However, the relationship between education level and death by suicide was less consistent among females. This sex disparity was also observed in the existing literature(31, 32) and thus, a better understanding of this sex disparity in the relationship between education level and suicide is warranted. Further, it is possible that other indicators of SES could act as moderators of the focal relationship investigated here, therefore, more research in this area needs to be conducted in order to draw conclusions on whether SES moderates the relationship between average amount of alcohol consumed per day and death by suicide.

The study findings should be interpreted in light of the following limitations. First, as indicated above, it was not possible to account for the complex survey design of the NHIS in the Aalen’s additive hazard models. However, when we compared the results of the Cox proportional hazards models adjusting for survey design with those not adjusting for survey design, similar findings were observed. Thus, we suspect that the impact of not adjusting for survey design on the effect estimates and our inferences based on the Aalen’s additive hazard models would be minimal. Second, previous studies have suggested that the spirits-suicide relationship may be stronger than that for beer and wine due to more rapid ethanol intake and intoxication and higher levels of aggression and emotional response associated with spirits consumption (33, 34). However, there is no beverage type information in the NHIS, and thus we were not able to conduct beverage-specific analyses or account for the different sizes and alcohol by volume percentages of beer, wine, and spirits in the calculation of alcohol g/day. Third, the data on average alcohol volume consumed per day arose from participants’ self-report at a single time point with an average follow-up time of nearly 11 years. Thus, reporting bias and changes in alcohol use over time may have introduced misclassification and underestimates the association between average alcohol volume consumed and death by suicide.

Despite the limitations, the current study represents the first step towards understanding the relationship between average alcohol volume consumed per day and suicide. It is evident that additional research is needed, including repeated measures of average alcohol consumption over the follow-up period. Only once additional research is carried out, will we gain full insight into whether the predisposing effects of alcohol on suicide mortality risk include average alcohol volume consumed per day, and at what level in particular.

## Data Availability

No data are available. Supporting data are not available due to ethical/legal restrictions.

## Contributors

SL conceptualized the study, interpreted the data, drafted the initial manuscript, reviewed, and revised the manuscript and prepared/formatted the manuscript for submission. YZ performed the statistical analyses and reviewed and revised the manuscript. CP conceptualized the study, coordinated, and supervised the data analyses, and critically reviewed the manuscript for important intellectual content. All authors approved the final manuscript as submitted and agree to be accountable for all aspects of the work.

## Funding

This work was supported by the National Institute on Alcohol Abuse and Alcoholism of the National Institutes of Health under grant award numbers R01AA028009. The content is solely the responsibility of the authors and does not necessarily represent the official views of the National Institutes of Health.

## Competing interests

No competing interests to declare.

## Data availability statement

No data are available. Supporting data are not available due to ethical/legal restrictions.

## Notes

### Competing Interest Statement

The authors have declared no competing interest.

### Author Declarations

All adult participants in the NHIS provided written informed consent. NHIS is approved by the Research Ethics Review Board of the National Center for Health Statistics and the U.S. Office of Management and Budget.

