## Supplemental Table 1 for "Evaluation of the risk relationship between average alcohol volume consumed and suicide: An analysis of mortality linked cohort data"

**Supplemental Material**

**Table S1.** Results of the hazards model^a^ testing the interaction between average alcohol volume consumed and level of education on death by suicide

|  | HR | 95% CI | p-value | Additional deaths per 100,000 person years | 95% CI | p-value |
| --- | --- | --- | --- | --- | --- | --- |
| Males |  |  |  |  |  |  |
| Level of education |  |  |  |  |  |  |
| High school diploma or less | **1.87** | **1.07, 3.26** | **0.029** | 8.6 | -0.6, 17.8 | 0.066 |
| Some college | 1.74 | 0.95, 3.21 | 0.075 | 10.5 | -0.4, 21.5 | 0.058 |
| Bachelor’s degree or higher | Ref | - | - | Ref | - | - |
| Alcohol use |  |  |  |  |  |  |
| Lifetime abstainer | Ref | - | - | Ref | - | - |
| Former drinker | 1.46 | 0.57, 3.73 | 0.433 | 4.3 | -13.3, 21.9 | 0.633 |
| (0, 20] g/day | 1.25 | 0.74, 2.12 | 0.399 | 0.9 | -6.9, 8.8 | 0.815 |
| (20, 40] g/day | 1.58 | 0.75, 3.33 | 0.230 | 4.7 | -9.2, 18.5 | 0.509 |
| (40, 60] g/day | 1.08 | 0.31, 3.82 | 0.905 | 0.9 | -22.8, 24.6 | 0.939 |
| >60 g/day | 1.75 | 0.48, 6.35 | 0.394 | 20.8 | -25.6, 67.2 | 0.379 |
| Interaction effects |  |  |  |  |  |  |
| High school diploma or less: Former drinker | 1.04 | 0.37, 2.92 | 0.945 | 8.2 | -13.6, 30.0 | 0.462 |
| Some college: Former drinker | 1.07 | 0.34, 3.35 | 0.909 | 8.9 | -17.0, 34.7 | 0.500 |
| High school diploma or less: (0, 20] g/day | 0.68 | 0.37, 1.25 | 0.212 | -0.7 | -11.2, 9.9 | 0.904 |
| Some college: (0, 20] g/day | 0.66 | 0.34, 1.28 | 0.220 | -4.9 | -17.4, 7.6 | 0.444 |
| High school diploma or less: (20, 40] g/day | 0.58 | 0.22, 1.53 | 0.272 | -6.7 | -24.7, 11.4 | 0.469 |
| Some college: (20, 40] g/day | 0.65 | 0.24, 1.79 | 0.405 | -4.1 | -24.9, 16.8 | 0.702 |
| High school diploma or less: (40, 60] g/day | 1.48 | 0.39, 5.68 | 0.567 | 22.2 | -11.3, 55.7 | 0.194 |
| Some college: (40, 60] g/day | 2.25 | 0.54, 9.35 | 0.266 | 39.5 | -5.4, 84.3 | 0.085 |
| High school diploma or less: >60 g/day | 0.6 | 0.13, 2.70 | 0.507 | -14.1 | -64.2, 36.1 | 0.582 |
| Some college: >60 g/day | 0.84 | 0.17, 4.03 | 0.827 | -7.5 | -64.4, 49.4 | 0.796 |
| Females |  |  |  |  |  |  |
| Level of education |  |  |  |  |  |  |
| High school diploma or less | 0.28 | 0.06, 1.20 | 0.085 | 0.1 | -3.0, 3.1 | 0.974 |
| Some college | 0.59 | 0.13, 2.67 | 0.494 | 2.2 | -1.6, 6.0 | 0.258 |
| Bachelor’s degree or higher | Ref | - | - | Ref | - | - |
| Alcohol use |  |  |  |  |  |  |
| Lifetime abstainer | Ref | - | - | Ref | - | - |
| Former drinker | 1.43 | 0.29, 7.12 | 0.666 | 12.2 | -1.7, 26.1 | 0.085 |
| (0, 20] g/day | 0.5 | 0.12, 2.08 | 0.339 | 1.3 | -2.2, 4.7 | 0.465 |
| >20 g/day | 0.8 | 0.14, 4.61 | 0.801 | 4.1 | -7.0, 15.3 | 0.468 |
| Interaction effects |  |  |  |  |  |  |
| High school diploma or less: Former drinker | 1.5 | 0.22, 10.35 | 0.680 | -7.3 | -22.5, 7.8 | 0.343 |
| Some college: Former drinker | 0.96 | 0.13, 6.92 | 0.970 | -9.6 | -26.1, 6.8 | 0.251 |
| High school diploma or less: (0, 20] g/day | 4.26 | 0.88, 20.53 | 0.071 | 0.6 | -3.5, 4.7 | 0.779 |
| Some college: (0, 20] g/day | 2.51 | 0.51, 12.36 | 0.258 | 0.7 | -4.1, 5.5 | 0.776 |
| High school diploma or less: >20 g/day | 1.28 | 0.12, 13.44 | 0.834 | -3.4 | -16.6, 9.8 | 0.612 |
| Some college: >20 g/day | 2.34 | 0.29, 18.90 | 0.424 | 10.3 | -9.4, 29.9 | 0.307 |

g: grams; HR: hazard ratio; Ref: reference category

^a^Adjusted for race/ethnicity, marital status, and survey year

*Note.* Bolded text signifies statistical significance at α=0.05.
